# Genistein effect on cognition in early Alzheimer’s disease patients. The GENIAL clinical trial

**DOI:** 10.1101/2022.06.01.22275832

**Authors:** José Viña, Joaquín Escudero, Miquel Baquero, JA Carbonell-Asíns, Francisco J. Tarazona-Santabalbina, Mónica Cebrián, José Enrique Muñoz, Encarnación Satorres, Juan Carlos Melendez, José Ferrer Rebolleda, Ma. del Puig Cózar Santiago, Jose Manuel Santabárbara Gomez, Mariona Jové, Reinald Pamplona, Consuelo Borrás

## Abstract

**Background:** Delaying the transition from minimal cognitive impairment to Alzheimer’s dementia is a major concern in Alzheimer’s disease (AD) therapeutics.

Pathological signs of AD occur years before the onset of clinical dementia. Thus, long-term therapeutic approaches, with safe, minimally invasive, and yet effective substances are recommended. There is a need to develop new drugs to delay Alzheimer’s dementia. We have taken a nutritional supplement approach with genistein, a chemically defined polyphenol that acts by multimodal specific mechanisms. Our group previously showed that genistein supplementation is effective to treat the double transgenic (APP/PS1) AD animal model.

**Methods:** In this double-blind, placebo-controlled, bicentric clinical trial we evaluated the effect of daily oral supplementation with 120 mg of genistein for 12 months on 24 early symptomatic Alzheimer’s patients. We used a battery of validated neurocognitive tests: Mini-Mental State Exam (MMSE), Memory Alteration Test (M@T) Clock-drawing test, Complutense Verbal Learning Test (TAVEC), Barcelona Test-Revised (TBR), and Rey Complex Figure Test.

**Results:** We report that genistein treatment results in a significant improvement in two of the tests used (dichotomized direct TAVEC, p=0.031; dichotomized delayed centil REY copy p=0.002 and a tendency to improve in all the rest of them.

The amyloid-beta deposition was analyzed using 18F-flutemetamol uptake which showed that genistein-treated patients did not increase their uptake in the anterior cingulate gyrus after treatment (p = 0.878) while placebo-treated did increase it (p=0.036) We did not observe significant changes in other brain areas studied

**Conclusions:** This study shows that genistein may have a role in therapeutics to delay the onset of Alzheimer’s dementia in patients with mild cognitive impairment. These encouraging results indicate that this should be followed up by a new study with more patients to further validate the conclusion that arises from this study.

**Trial registration:** NCT01982578

## BACKGROUND

Alzheimer’s disease (AD) treatments, especially those aiming at changing the course of the disease, must start decades before the onset of full-blown dementia because the disease’s histopathological hallmarks can be observed many years before the clinical symptoms start [1].

Hence, treatments aiming at changing the course of the disease over decades should be as innocuous, non-invasive, and inexpensive as possible.

Some treatments, especially those in which intravenous injections of Amyloid β (Aβ) - directed antibodies have shown some promising results [2]. However, these antibodies must be administered parentally for prolonged periods. Aducanumab was recently approved by the FDA as a treatment for Alzheimer’s [3].

Genistein is a chemically defined phytoestrogen present in soya and has been reported to have beneficial properties on age-related diseases such as neurodegenerative [4] and cardiovascular diseases [5] or cancer [6]. It is a multimodal agent: it acts as an antioxidant, anti-inflammatory, and anti-Aβ as well as an autophagy promoter [7].

Our previous studies using genistein demonstrated that it is useful to reverse the loss of cognition and pathological hallmarks in animal models of Alzheimer’s disease [4] by targeting specific pathophysiological and signaling mechanisms involved in the disease [7, 8]. These animal results required translation into humans.

Here we report the results of the GENIAL clinical trial (NCT01982578) with genistein vs placebo on patients suffering from mild cognitive impairment. The primary outcome was to determine differences in the deposition of Aβ in the brain after one year of treatment. The secondary outcome was to analyse the effect on the progressive loss of cognition after six months and one year of treatment. Our results indicate that early AD patients treated for one year with genistein, show a lower amyloid *β* deposition as determined by Flutemetamol uptake in a specific brain area. They also lose less cognition than controls in two of the tests used. A tendency to improve was observed in all the rest of the tests. These results open the possibility of carrying out a phase two clinical trial with a larger number of patients.

## METHODS

### Trial oversight

INC-GEN-2013-01 (GENIAL) is a bicentric, randomized, double-blind, placebo-controlled phase 2 clinical study that assessed the efficacy of genistein in patients with early Alzheimer’s disease. The study was conducted in 2 public hospitals in the city of Valencia, Spain: The Department of Neurology of Hospital General and Hospital La Fe according to the protocol (see https://clinicaltrials.gov/ct2/show/NCT01982578) and with the consensus ethics principles derived from international ethics guidelines, including the Declaration of Helsinki and the Council for International Organizations of Medical Sciences International Ethical Guidelines. Study participants provided written informed consent. Patients were visited 3,6,9 and 12 months after the start of the trial. Cognition was evaluated at baseline, at six months, and at the end of the study. Patients were recruited at the Neurology departments of Hospital General and Hospital La Fe, both in Valencia, Spain.

The trial director, Professor José Viña, designed the trial, which was funded by a grant by the Spanish Government (SAF2016-75508-R from the Spanish Ministry of Economy and Competitivity). The authors vouch for the accuracy and completeness of the data, and the fidelity of the study to the protocol.

Figure 1 shows that we recruited 32 patients that were eligible for the study. Of those, 27 underwent blinded randomization then 14 were assigned to the genistein and 13 to the placebo. Of the 14 patients that were assigned genistein, one withdrew and hence 13 patients completed the trial. Of the 13 that were assigned placebo, two discontinued the treatment, because they had adverse events and 11 completed the trial.

**Figure 1.**
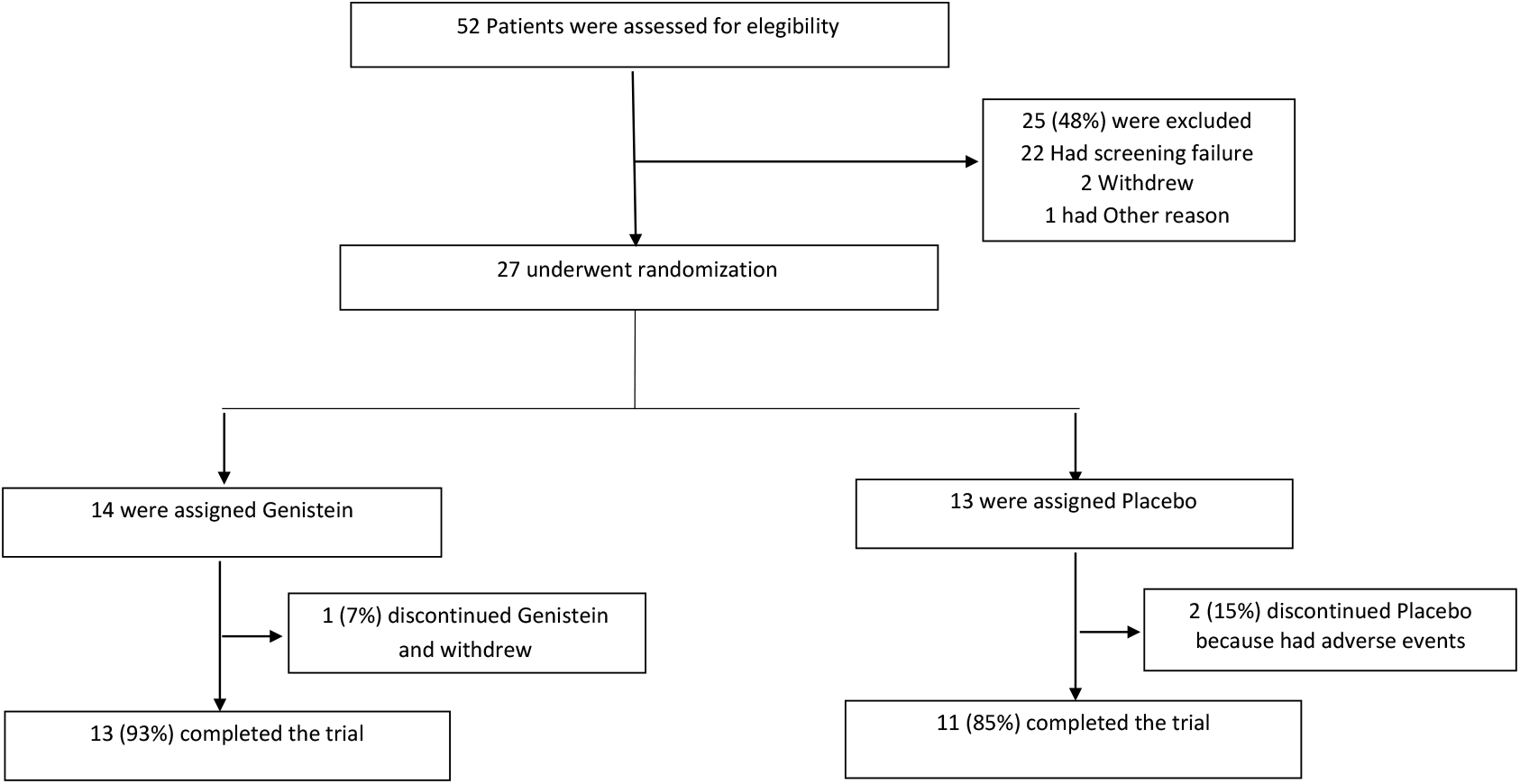
Enrollment, randomization, and clinical completion.

### Eligibility criteria

The study included patients 54 to 75 years of age who met Dubois (IRW_2) criteria for cognitive impairment compatible with prodromal Alzheimer’s disease or mild dementia of Alzheimer’s type [9] and had a Mini-Mental State Examination (MMSE) score ≥ 24 (scores range from 0 to 30, with higher scores indicating better mental performance). Screening procedures included the MMSE, ApoE determination, positron emission tomography (PET) with 18F-Flutemetamol to confirm Amyloid β deposition, and nuclear magnetic resonance to exclude vascular dementias. PET scans were reviewed at a centralized PET imaging facility for assessment of eligibility. Patients were required to have Flutemetamol PET scans, and the scans were quantitatively evaluated for estimation of an Amyloid β standardized uptake value ratio (SUVR) according to published methods [10–12]. Three patients with a negative amyloid PET were excluded.

### Interventions

Patients who met the eligibility criteria were randomly assigned in a 1:1 ratio to receive either 1 capsule of genistein (60 mg) or a placebo, administered orally twice per day for up to 12 months. The capsules were purchased from Zambon Spain who commercialized them under the brand name Fisiogen®. Capsules then were prepared by the Pharmacy Department of Hospital Clínico Universitario of Valencia so that the capsules were equal in shape and size for both groups Randomization was performed with SPSS IBM statistics (version 22.0) for Windows and the Randomization list was archived in the Pharmacy file. Unblinding was not performed during the clinical trial. A seed value has been used to reproduce the results. Blocks of four patients were used (2 active treatments and 2 placebos per block to ensure the same number of patients in each arm.) Two dispensing visits were performed on day 1 and day 180. The patients returned non-used medication on day 360. Compliance was verified and recorded in the CRD by the investigator.

### Safety Assessments

Safety assessments were performed by site investigators who were unaware of the study group assignments. Safety outcomes included spontaneously reported adverse events, clinical laboratory test results, vital signs and body-weight measurements, findings on 12-lead electrocardiography, and physical and neurologic examinations. Details regarding safety follow-up visits (which are ongoing) are provided in the study protocol.

### Plasma genistein level determination

Genistein was extracted, processed, and purified from plasma samples based on a previously described method [13, 14]. Briefly, 100 µl of acetonitrile were added to 100 µl of plasma with labeled internal standard (d4 genistein 0.1µM, Cluzeau Info Labo, Sainte Foy la Grande, France), vortexed for 1 min, and centrifugated at 12.000 g for 2 min for protein precipitation. The supernatants were collected and subjected to enzymatic hydrolysis by adding 2.3 units of sulfatase/glucuronidase H1 (Sigma Aldrich, St. Louis, MO) in 25 mM sodium citrate pH5. The resulting solutions were then purified using a solid-phase extraction (SPE) method. We used Bond Elut C18 96 round-well plate (Agilent Technologies, Barcelona, Spain) under reduced pressure. Conditioning of the cartridge was achieved with 1 mL acetonitrile followed by 1 mL of 25 mM sodium citrate buffer at pH5. Then, enzyme-treated plasma samples were applied to the plate and subsequently washed with 1 mL of water followed by 1mL of 10% aqueous methanol. Analytes were eluted with 6 × 200 µL aliquots of 5% formic acid in acetonitrile. The eluate was dried in a SpeedVac (Fisher Scientific, Madrid, España) and resuspended in 50 µl of water 01.% formic acid/acetonitrile (60/40, v/v). Final extracts were filtered in an Eppendorf UltraFree 5 kDa filter (Millipore, Billerica, MA).

For detection and quantification of genistein, 15µl of filtered final extracts were submitted to ultraperformance liquid chromatography (UPLC) using a Waters Acquity ultraperformance liquid chromatography system (Waters, Milford, MA), equipped with a binary pump system (Waters, Milford, MA) using an Acquity UPLC BEH C18 column (1.7µm, 2.1 * 50mm) (Waters, Milford, MA) with a binary mobile phase and coupled to a TQD mass spectrometer (Waters, Milford, MA) using a Z-spray ESI source operating in positive mode, as previously described [14]. Data acquisition was carried out with MassLynx v 4.1 software. Optimized MRM conditions are: 270.9 ->214.9 for genistein and 275.0 ->218.9 for d4 genistein. The amounts of product were expressed as nmol of genistein being adjusted from the deuterated internal standard.

### Amyloid-beta assessment by positron emission tomography (PET)

Amyloid-*β* PET was performed using a Gemini TF PET/CT scanner (Philips Medical Systems, the Netherlands). Amyloid-PET images were acquired 90 minutes after the intravenous administration of 18F-Flutemetamol tracer (185 MBq).

All scans were initially read by an expert nuclear medicine physician (MPC, read 1). In addition, the scans were reread for this study by JFR (read 2), while being both blinded to the results of other visual reads, cerebrospinal fluid (CSF), and arm of the study.

Amyloid-*β* PET status (positive or negative) was determined by a majority visual read agreement of two readers and a consensus position after reviewing together the images in studies with conflicting initial results.

Additionally, quantification of brain regions was obtained using the MIM Neuro application (MIM Software Inc., Cleveland, OH, USA). SUV Ratio (the ratio of the average value for each brain region to the average value for the normalization structure (cerebellum) was used for quantifying the Amyloid plaque burden in the brain confronting each patient’s results to the normal MIM Flutemetamol database.

### Efficacy Outcomes

The primary outcome was the change from baseline to 360 days in brain β Amyloid levels by SUVR determination, defined as the difference between the values at the beginning and end of follow-up in each patient.

Secondary outcomes were changes in a battery of different neuropsychological tests, which were administered to all patients before randomization, at 180 days of treatment, and after 360 days of treatment. The following tests were administered to all patients: Mini-Mental State Exam (MMSE) [15], which is a widely used test of cognitive function among older adults; it includes tests of orientation, attention, memory, language, and visual-spatial skills (scores range from 0 to 30, with higher scores indicating better mental performance). Memory Alteration Test (M@T) [16] is a screening test for AD that assesses verbal episodic and semantic memory. It has five subtests: encoding, temporal orientation, semantic memory, free recall, and cued recall. The maximum M@T score is 50 points and a score below 37 is an optimal cut-off score. Clock-drawing test is used to assess a patient’s cognitive abilities and detect possible cognitive impairment. Based on Rouleau et al. [17], the maximum score is 10 points on each of the three subscales. Lower scores indicate greater deterioration. The Complutense Verbal Learning Test (TAVEC) [18] consists of a list of 16 words that are presented to the subject five times to evaluate different memory and learning processes. Each trial is scored from 0 to 16, with a maximum of 80 for the Total TAVEC score. For the measurement of the Delayed TAVEC, 20 min after the immediate recall, the patient must remember the previously read list. The Barcelona Test-Revised (TBR) [19] consists of two subtests, the semantic verbal fluency (name animals for one minute) and the phonological verbal fluency (words that begin with the letter “p” for three minutes). These tests were administered to assess language ability and observe the capacity for accessing and evoking elements from the lexical and semantic warehouse. The Rey Complex Figure Test [20] allows the assessment of a wide variety of cognitive processes, such as memory and executive functions, organizing and planning, and visuo-constructive and spatial skills. The patient must carefully reproduce a complex geometric drawing (Copy Rey) and later (3 min) reproduce it from memory. Each of these attempts is scored directly on a scale ranging from a minimum of 0 to a maximum of 36 points; in addition, the centile score was used.

### Statistical analysis

Qualitative variables were described using frequencies and percentages while quantitative variables with mean and standard deviation. Normality of the continuous variables was assessed according to the Shapiro-Wilks test and if their normal distribution was not confirmed, they were described using the median and interquartile range. For quantitative variables, the mean comparison was carried out using the t-Student test if their normality assumption held true otherwise, the Mann-Whitney test. For qualitative variables, a comparison of percentages between groups was carried out with Fisher’s exact test for dichotomous variables or the chi-square test for variables with more than two categories. Changes in genistein level between treatment groups after twelve months were evaluated using a linear mixed model with individuals as a random effect, time (baseline versus 12 months), treatment (genistein versus placebo), and interaction between these two as fixed effects. The same model was also used to study neuropsychological tests and CADS. Delayed Centil REY copy was recoded into “Low” if the value was lower than 1 and “High” otherwise. The effect of treatment effect was then studied using a generalized mixed model of the binomial family with individual id as a random effect, time (baseline versus 12 months), treatment (genistein versus placebo), and interaction between these two as fixed effects. Direct TAVEC was dichotomized into worsening/no-change versus improvement after 12 months Treatment effect was then studied using Fisher’s exact test.

The software used for all analysis is R in its 3.6.1 version (R Core Team, 2019). All mixed model p-values were adjusted with the Tukey method for comparing a family of 4 estimates. The cutoff for significance was set to α = 0.05.

## RESULTS

### Patient characteristics

Table 1 shows the characteristics of the patients. Age, sex, education levels, and *APOE* genotype were not statistically different between control and genistein-treated groups.

**Table 1.**
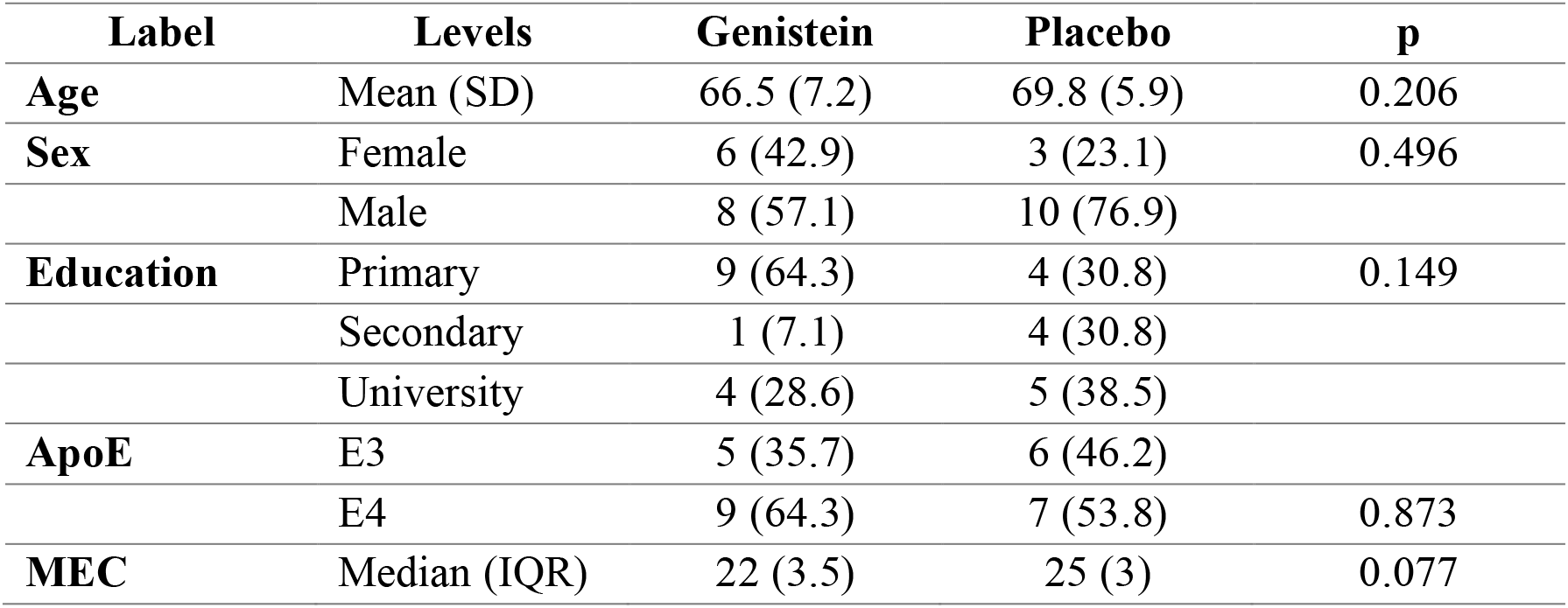
Characteristics of the participants at baseline.

### Plasma genistein concentrations

To test the effect of administration of genistein on plasma levels, we determined genistein (both free and conjugated) in the plasma of the patients. To have the whole pool of genistein present in plasma samples we enzymatically treated the samples before extraction and quantification (see Methods). Figure 2 shows that individuals taking a placebo showed a very low level of genistein at both the beginning and the end of the study. By contrast, individuals who took genistein showed a low level of genistein at the beginning of the study, indistinguishable from those taking placebo, but a significant increase in genistein at the end of the study. The differences in plasma genistein between individuals taking a placebo and those taking genistein at the end of the study were significant as they were also between individuals in the genistein treatment group before and after treatment.

**Figure 2.**
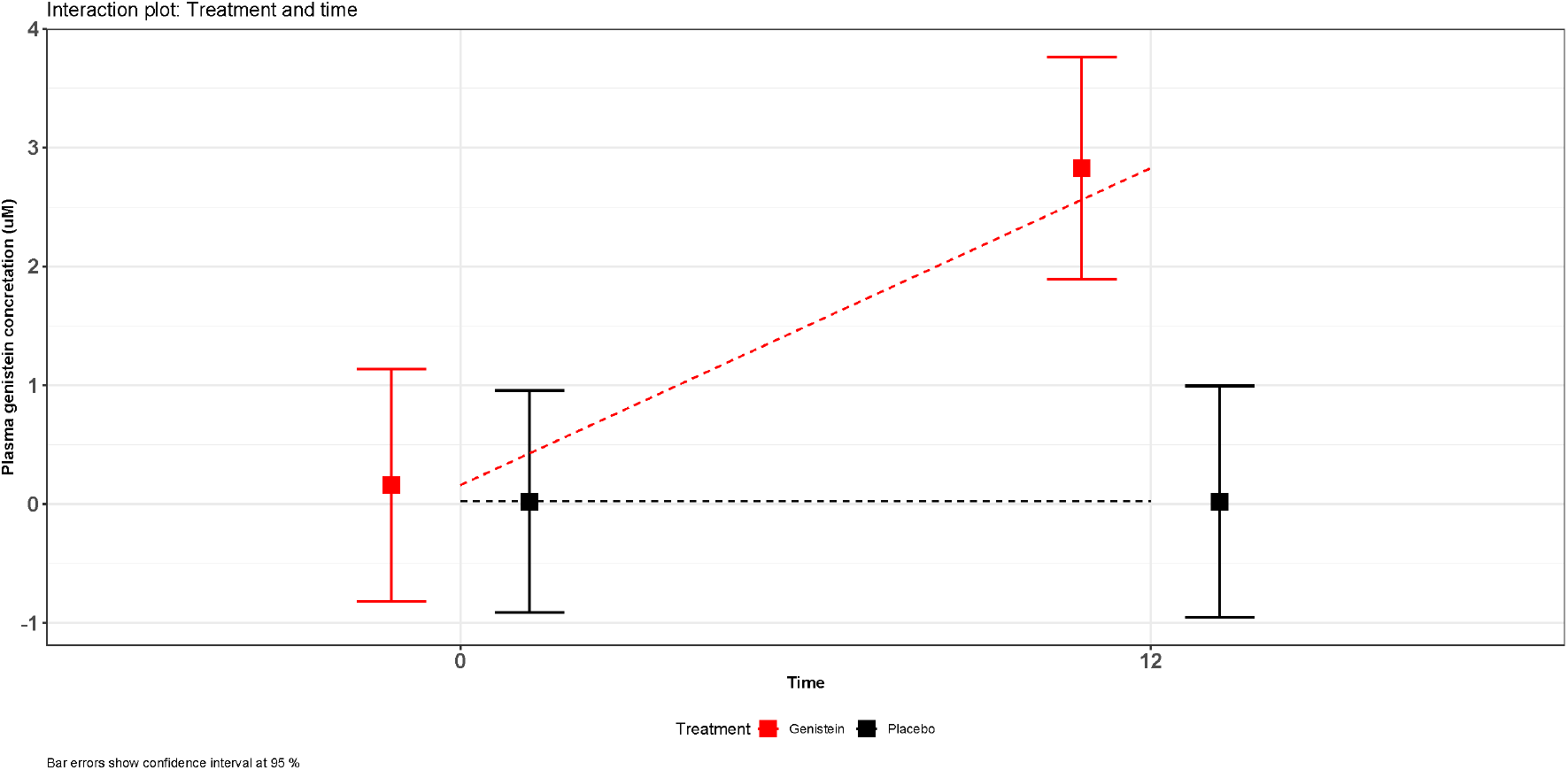
Plasma genistein concentration at baseline and after 12 months of genistein treatment. Boxes show adjusted means while confidence intervals are represented with error bars of n=12-13 individuals.

### Effect of genistein treatment on flutemetamol uptake by different brain areas

This was the primary outcome of the trial. We determined the standardized uptake value ratio (SUVR) which is the quantitative determination of beta-amyloid deposits as measured by flutemetamol PET. It consists of quantifying the standard uptake value in different cortical regions of the brain versus a standard uptake value of a reference area (cerebellum).

The values obtained were not statistically different between the placebo and genistein groups in the whole brain or specific areas, except for the anterior cingulate gyrus. 18F-flutemetamol uptake in this area rose in placebo-treated patients (p=0.036) but did not increase in genistein-treated ones (p = 0.878).

### Effect of genistein treatment on individual cognitive tests/Clinical outcomes

We determined the values of each one of the cognitive tests analyzed at the beginning of the study and 6 and 12 months after treatment with genistein or placebo. We report here the results at 12 months of treatment. Results at 6 months were non-significant (results not shown).

Of all the individual tests studied, only total dichotomized TAVEC and Centil REY Delayed copy showed a statistically significant improvement in the genistein group compared with the placebo group see Figure 3A (p=0,03 for Direct TAVEC) and Figure 3B for dichotomized Centil score REY Delayed).

**Figure 3.**
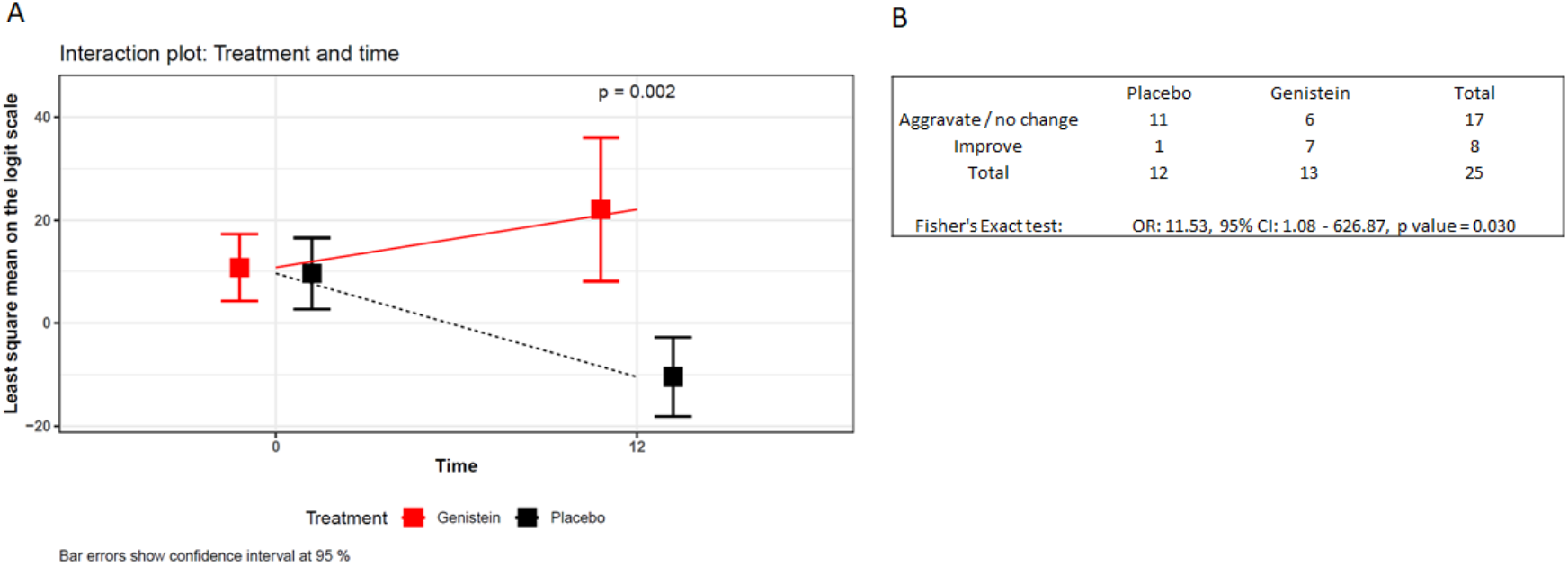
Genistein effects on cognitive tests. Interaction plot between treatment and time for dichotomized TAVEC (A). The contingency table with dichotomized REY copy values and the p-value used has been calculated using Fisher’s exact test (B).

Moreover, in all the tests analyzed, there was a tendency to improve in genistein-treated patients. The results are shown in the supplementary material (see Figure S1), and they are commented on in the discussion section.

### Adverse effects

There were no reported death or serious adverse events in the genistein group or the placebo group. In the placebo group, two of the 13 patients withdrew from the trial, one was institutionalized in a residence because of a bad progression of the AD, and the other one due to gastrointestinal problems that the patient associated with the investigational treatment. In the genistein group, one of the 14 patients withdrew from the trial because he referred to intestinal problems (increased depositions number).

## DISCUSSION

This trial is based on our previous experience in which genistein, a soy isoflavone, is effective in significantly improving the cognition of the APP/PS1 Alzheimer’s mice model [4]. Genistein acts by defined mechanisms essentially: lowering amyloid levels in the brain, improving antioxidant status, and lowering brain inflammation [7, 8].

The importance of genistein as a potential tool against Alzheimer’s” has been reviewed [21]. The possible concern regarding the low bioavailability of genistein is minimized by the fact that very low plasma concentrations may be sufficient because many of the actions of genistein are receptor-mediated. Nevertheless, it is very important to determine the plasma level of genistein when using it for clinical purposes [22]. We show here that indeed they are higher in the genistein-treated patient than in controls. We have studied the effect of a nanodrug containing genistein and compared it with oral free genistein and have observed that the favorable effects are very similar in both cases [23].

The primary outcome was the determination of the level of amyloid-beta plaques. We determined it by analyzing the flutemetamol uptake in the brain by positron emission tomography. Global analysis of the brains of patients did not show any significant difference before or after twelve months of treatment. Analysis of different brain areas only showed differences in the anterior cingulate gyrus. In this particular area, patients treated with genistein accumulated fewer amyloid deposits after 12 months of treatment than patients treated with placebo. In our previous animal studies, we found that genistein lowers the amyloid-beta load in the whole brain. This was further confirmed *in vivo* by positron emission tomography [4]. Moreover, Bagheri et al. already showed in 2012 that treatment with genistein prevents the formation of amyloid-beta aggregates in the lateral blade of the dentate gyrus region of the brain of rats previously injected with a solution of amyloid-beta [24].

Nevertheless, a recent paper underpinned the variability of imaging results and, due to the small sample size, proposed that significant results can only be obtained if thousands of participants are recruited. This is an obvious limitation of these techniques as thousands of patients are seldom recruited in the usual clinical trials [25].

The secondary outcomes proposed dealt with cognition. All the nine parameters measured showed a tendency of improvement after treatment with genistein when compared with placebo. Of these, two reached significant differences, the total TAVEC test, and the Centil score Rey delayed. The TAVEC test provides information on immediate recalling and verbal learning. The Rey delayed test indicates the capacity of copying and then retrieving a complex geometrical figure. This gives information on executive functions, attention, and immediate visual memory.

The favorable effect of genistein on experimental animals was observed by Bagheri in an acute model of experimental Alzheimer’s disease (micro-injection of αβ in mice) [24] and by ourselves (using the APP-PSEN1 Alzheimer’s transgenic model) [4]. Results were encouraging and thus this clinical trial was started in 2013. Other groups have also tested the effects of soy isoflavones in AD patients. Gleason et al. did not observe significant effects on cognition after soya isoflavones administration, but they administered the soy isoflavones only for six months [26]. This agrees with our results, as we did not find improvements at six months of treatment but did find them after one year. This time-cause effect underpins the importance of relatively long-term treatments to prevent or delay the transition to full-blown dementia in Alzheimer’s minimal cognitive impairment patients.

Because all the tests resulted in a tendency to improve, even though we only found a statistically significant improvement in two of them in the genistein-treated patients compared with the placebo group, we performed a combined Alzheimer’s disease score calculated in two steps: first, the difference between the various scores at twelve months minus the value at the beginning of the study was calculated and in a second step all the variables were added together. Therefore, lower scores imply a worse cognitive state. The minimum and maximum in our cohort were −156.2 and 11 respectively. Using this score genistein-treated patients performed significantly better than placebos (genistein-treated patients had −14.83 (24.18) points whereas placebos had −55.89 (53.28), the difference is 41.06 (95% CI: 5.13-76.99, p=0.028). This may be explained because each one, without exception, of the tests measured was better in patients treated with genistein than in those with placebo, albeit only two modified tests showed statistical significance on their own.

### Strengths and limitations of this study

A strength of this study is that we propose treatment with a safe, widely used substance whose multimodal mechanisms of action [8] have been defined in animal models. Genistein acts by lowering inflammation [27], lowering Aβ deposition in the brain [4], and increasing antioxidant defenses in experimental animals [5]. A limitation of this study is the low number of patients and the fact that the treatment period was of only one year. Thus, this must be considered a pilot study. The encouraging results indicate that this should be followed up by a new study with more patients to confirm or disprove the conclusion we reach here.

## CONCLUSIONS

This study shows that genistein may have a role in therapeutics to delay the onset of Alzheimer’s dementia in patients with mild cognitive impairment. These encouraging results indicate that this should be followed up by a new study with more patients to further validate the conclusion that arises from this study.

## Data Availability

All the relevant data are included in the manuscript. Other data produced in the presente study are available upon reasonable request

## LIST OF ABBREVIATIONS

(AD): Alzheimer’s disease
(MMSE): Mini-Mental State Exam
(M@T): Memory Alteration Test
(TAVEC): Complutense Verbal Learning Test
(TBR): Barcelona Test-Revised
(SUVR): standardized uptake value ratio
(Aβ): Amyloid β
(PET): positron emission tomography.

## DECLARATIONS

### Ethics approval and consent to participate

The study was conducted across 2 sites in the city of Valencia, Spain, at the Department of Neurology of Hospital General and Hospital La Fe according to the protocol (see https://clinicaltrials.gov/ct2/show/NCT01982578) and with the consensus ethics principles derived from international ethics guidelines, including the Declaration of Helsinki and the Council for International Organizations of Medical Sciences International Ethical Guidelines. Study participants provided written informed consent. The trial director, Professor José Viña, designed the trial, which was funded by a grant by the Spanish Government (SAF2016-75508-R from the Spanish Ministry of Economy and Competitivity). The authors vouch for the accuracy and completeness of the data, and the fidelity of the study to the protocol.

### Consent for publication

All the authors have given their consent for publication

### Availability of data and materials

The data and materials are available at clinicaltrials.gov under the registration number NCT01982578 (https://clinicaltrials.gov/ct2/show/NCT01982578).

### Competing interests

This was supported exclusively by public funds in Spain and was performed in two public hospitals in Valencia. No private industry support must be disclosed. Genistein-containing pills were bought from commercially available sources that were distributed in the chemist shop system in Spain.

### Funding

This work was supported by the following grants: CB16/10/00435 (CIBERFES) from Instituto de Salud Carlos III, (PID2019-110906RB-I00/ AEI / 10.13039/501100011033) and RED2018-102576-T from the Spanish Ministry of Innovation and Science, PROMETEO/2019/097 from “Consellería de Innovación, Universidades, Ciencia y Sociedad Digital de la Generalitat Valenciana” and EU Funded H2020-DIABFRAIL-LATAM (Ref: 825546), European Joint Programming Initiative “A Healthy Diet for a Healthy Life” (JPI HDHL) and of the ERA-NET Cofund ERA-HDHL (GA N° 696295 of the EU Horizon 2020 Research and Innovation Programme) and Fundación Ramón Areces y Fundación Soria Melguizo. to J.V. and Grant PID2020-113839RB-I00 funded by MCIN/AEI/ 10.13039/501100011033, PCIN-2017-117 of the Ministry of Economy and Competitiveness, and the EU Joint Programming Initiative ‘A Healthy Diet for a Healthy Life’ (JPI HDHL INTIMIC-085) to CB. We also acknowledge funding from the Spanish Ministry of Science, Innovation, and Universities (RTI2018-099200-B-I00), the Generalitat of Catalonia, Agency for management of University and Research Grants (2017SGR696) and the Department of Health (SLT002/16/00250) to RP. Part of the equipment employed in this work has been funded by Generalitat Valenciana and co-financed with ERDF funds (OP ERDF of Comunitat Valenciana 2014-2020). M.J. is a “Serra Hunter” Fellow.

### Authors’ contributions

J.V. conceived the trial, designed the protocol, interpreted the data, drafted and revised the manuscript, J.E., M.B., E.S., and J.C.M. selected the patients, performed the cognitive analysis, and interpreted the data, M.C. acted as a project manager and helped draft the manuscript, J.A.C.-A., performed the statistical analysis., J.E.M. prepared the drugs, J.F.R, M.P.C.-S. and J.M.S.-G. performed the PET analysis, M.J and R.P. determined plasma genistein levels, and C.B. interpreted the data and drafted and revised the manuscript.

## Acknowledgments

We thank Mrs. Marilyn Noyes for her kind help in reviewing the English style of the manuscript

## Notes

### Competing Interest Statement

The authors have declared no competing interest.

### Clinical Trial

NCT01982578

### Author Declarations

Comite Etico de Investigacion Clinica. Hospital Clinico Universitario de Valencia at the Oct, 24, 2013 meeting.

