## Supplementary figures and images for "Genistein effect on cognition in early Alzheimer’s disease patients. The GENIAL clinical trial"

### Supplementary Figure 1

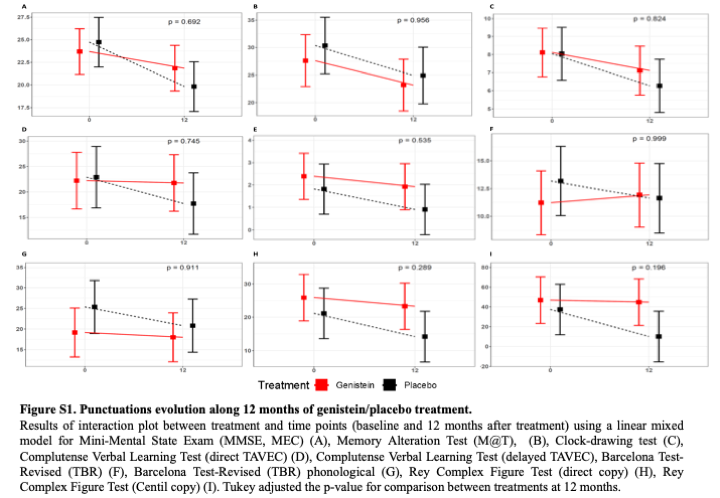
